# A pilot genome-wide association and gene-level prioritization analysis of ischemic heart disease with co-occurring arterial hypertension in a Kazakh cohort

**DOI:** 10.64898/2026.03.19.26348868

**Authors:** Liliya Skvortsova, Kanagat Yergali, Assel Zhaxylykova, Mamura Begmanova, Almagul Mansharipova

## Abstract

Genome-wide association studies (GWAS) of ischemic heart disease (IHD) remain underrepresented in Central Asian populations. We conducted a pilot GWAS of IHD with co-occurring arterial hypertension in a Kazakh cohort to identify candidate loci for future replication.

The study included 470 Kazakh individuals, comprising of 240 cases and 230 controls, genotyped by the Illumina Infinium Global Screening Array-24 v 3.0. Association testing was performed using logistic regression under an additive genetic model adjusted for age, sex and the first ten principal components. A total of 325 976 autosomal variants passed QC and were tested in the final association analysis. Bonferroni-corrected significance threshold was P < 1.53 × 10^-7^. No variant reached the significance threshold. The strongest SNP association was observed for rs1511924 at chromosome 16 mapped to *ZFHX3* gene (OR = 0.49, 95% CI = 0.37–0.66; P = 1.00 × 10^-6^). As secondary gene level analyses, we performed conventional MAGMA using GWAS derived SNPs p-values and KGWAS-informed MAGMA using KGWAS derived SNPs scores followed by MAGMA genes aggregation. Conventional MAGMA did not identify significant genes above the Bonferroni corrected threshold. KGWAS-informed MAGMA prioritized *TLE1* and *SMCO4* genes above the Bonferroni corrected threshold for the in-sample Kazakh LD reference panel. *CSMD1* gene was prioritized as top ranked but sub-threshold signal for the in-sample Kazakh LD reference panel.

This pilot study did not detect Bonferroni-significant SNP associations for IHD with co-occurring arterial hypertension in the Kazakh cohort but it prioritized candidate gene level signals using downstream conventional and knowledge-graph-informed analyses. The results highlight both the potential and the interpretational limitations of applying knowledge-graph-informed prioritization methods in small underrepresented populations and require replication in larger Central Asian cohorts and further validation.

## Introduction

Ischemic heart disease (IHD) remains one of the leading causes of death, disability and mortality worldwide (Shi et al., 2025). Recent analysis of World Health Organization mortality data from 105 countries shows that, despite the decline in overall and standardized mortality rates from IHD since 2000, the overall burden remains substantial. Moreover, mortality rate trends differ significantly by sex and region, with declines in Europe, North America and Oceania but stagnating or rising rates across much of Asia, Africa and Central and South America. Within this global pattern, Kazakhstan, a Central Asian country, was identified as having relatively high but gradually declining age-standardized mortality rates (ASMRs) (Mukasheva et al., 2022; Jaiswal et al., 2025). Nevertheless, cardiovascular diseases and IHD remain the leading cause of death in Kazakhstan. Recent, national statistics show that IHD mortality have reduced between 2011 and 2019 but is still substantial in terms of disability-adjusted life years (DALYs) and has even increased again in 2020–2021(Mukasheva et al., 2022) (Zhakhina et al., 2023). This increase in mortality coincided with the COVID-19 pandemic, which significantly disrupted the mortality structure and marked by an increase in the number and severity of cardiovascular events, particularly in younger adults (Kosherbayeva et al., 2024) (Junusbekova et al., 2023). In this context of high and unstable IHD burden worldwide and in Kazakhstan, a more comprehensive characterization of IHD determinants is essential in a setting of rapidly evolving epidemiological and lifestyle patterns. Clinically, IHD often manifests later in life and can be regarded as the result of cumulative exposure to modifiable environmental risk factors such as diet-related obesity, smoking status, physical inactivity, acting over decades on a background of genetic predisposition. Thus, even as lifestyle and environmental exposures continue to change, genetic susceptibility remains a crucial, relatively stable component of IHD risk that requires careful investigation. Family history of premature IHD is a strong, independent risk factor and twin and family studies suggest a substantial heritable component to IHD (Ozaki and Tanaka, 2016) (Miyazawa and Ito, 2021).

Genome-wide association studies (GWAS) identified hundreds of loci associated with IHD and myocardial infarction (MI), notably through large consortia such as CARDIoGRAMplusC4D and subsequent meta-analyses integrating UK Biobank and other cohorts (A comprehensive 1000 Genomes–based genome-wide association meta-analysis of coronary artery disease, 2015) (Nelson et al., 2015) (Tcheandjieu et al., 2022)(Kessler and Schunkert, 2021). These studies have uncovered novel genetic loci and biological pathways highlighted the polygenic nature of IHD. These findings have enabled the development of polygenic risk scores and the identification of new therapeutic targets (Miyazawa and Ito, 2021)(Matsunaga et al., 2020). More recent multi-ethnic GWAS have begun to extend these findings to genetically diverse populations, but important gaps remain. For example, to date, most IHD GWAS have been conducted in populations of European ancestry with more limited representation of East and South Asian cohorts and virtually no genome-wide data on IHD from Central Asia. Genome-wide data and linkage disequilibrium patterns for Central Asian populations are largely absent from widely used public reference panels. Current resources such as the 1000 Genome project and gnomAD include no or very few individuals from Central Asia, and several authors have highlighted this region as a major gap in the current population-genetic resources (Seidualy et al., 2020) (Karczewski et al., 2020) (Fatumo et al., 2022). At the same time, several whole-genome sequencing projects involving healthy Kazakh individuals have begun to characterize the genetic architecture of this population and provide initial reference data for Kazakhs (Akilzhanova et al., 2014) (Kairov et al., 2021) (Kairov et al., 2022). These studies also indicate that Kazakhs and other Central Asian groups exhibit a characteristic admixture of European and Asian ancestries. Substantial genetic heterogeneity underscoring the need for population-specific genomic data from this region (Narasimhan et al., 2019).

Against this background of limited population-wide genomic resources, disease-focused genetic research on IHD in Kazakhstan has also remained relatively sparse. In Kazakhstan, genetic associative research of IHD and other cardiovascular diseases has so far focused mainly on candidate variants. The variants included polymorphisms in the renin-angiotensin system (Baĭtasova and Rysmendiev, 2002) (Aitkhozhina and Lyudvikova, 2003), coagulation and inflammatory genes, 9p21.3 (*CDKN2B-AS1/ANRIL*) (Myngbay and Alibekov, 2023) (Aitkaliyev et al., 2024), loci related to coronary restenosis (Taizhanova et al., 2020) (Taizhanova et al., 2023), coronary artery disease in type 2 diabetes and cardiac autonomic neuropathy (Shakhanova et al., 2020) (Bekenova et al., 2024) (Aitkaliyev et al., 2024), hypertension (Muratbekova et al., 2025), stroke (Svyatova et al., 2023), as well as oxidative stress-related loci. In our previous case-control study, we investigated the *GCLM* -588C/T and *GCLC* -129T/C promoter polymorphisms in relation to IHD development in the population of Kazakhstan (Skvortsova et al., 2017). Despite the fact that these studies focused on a limited set of loci and do not capture the genome-wide architecture of cardiovascular diseases, they have provided initial insights into IHD and related cardiovascular phenotypes in this underrepresented Kazakh population. Thus, there is a clear need for more comprehensive, genome-wide approaches − including GWAS and next-generation sequencing (NGS), to better characterize the genetic landscape of IHD in Kazakh individuals. Although NGS-based approaches can yield detailed information at the level of whole genomes and exomes, their high cost currently restricts their use in Kazakhstan to relatively small numbers of samples. In contrast, GWAS using high-density SNP arrays offer a more feasible next step for population-scale studies in these settings, enabling the systematic identification of common risk variants for IHD in Kazakhs. Therefore, we conducted a pilot GWAS of IHD with co-occurring arterial hypertension in a population of ethnic Kazakhs to identify genetic variants associated with the combined phenotype of IHD and hypertension, (2) to examine whether these loci previously implicated in IHD or hypertension in other ancestries and (3) to explore potential population-specific signals that warrant replication in larger cohorts.

## Materials and Methods

### Ethics approval, biosamples and consent to participate

The study used a combination of archived and prospectively collected biosamples. A subset of peripheral blood samples had been collected previously, processed and stored as frozen archival material. The use of these archived samples for genetic research was approved by the Local Ethics Committee of Kazakh-Russian Medical University, Almaty, Kazakhstan (Approval No. 36, 1 May 2016). Archived samples and/or associated clinical records were accessed for research purposes on 3 February 2025. Additional peripheral blood samples were collected prospectively within the present project from 5 January 2024 to 30 August 2025 following approval by the Local Ethics Committee of the Institute of Genetics and Physiology, Almaty, Kazakhstan (Protocol No. 3, 30 October 2023; excerpt letter No. 13-368, 17 November 2023). Although the approved project title refers to next-generation sequencing, the approved study plan also included SNP-array genotyping, and the analyses reported here were conducted within the scope of the approved protocol.

The study was conducted in accordance with the Declaration of Helsinki and applicable national regulations. Written informed consent was obtained from all participants prior to enrollment and blood sample collection; for the archived samples, the use of previously collected samples was authorized under the ethics approval and applicable consent procedures in place at the time of collection.

### Study subjects

The study was conducted at City Clinical Hospital No. 1 (Almaty, Kazakhstan; Department of Cardiology) and Kazakh-Russian Medical University. Patients with a combined phenotype defined as IHD with co-occurring arterial hypertension (hereafter referred to as IHD – HTN co-occurrence) were recruited for the study. Peripheral blood samples were collected from 240 patients with clinically documented of IHD and arterial hypertension including stable angina pectoris (Canadian Cardiovascular Society classes I – IV and Association of Cardiologists of the Republic of Kazakhstan [<u>16</u>]), chronic heart failure (New York Heart Association classes II-III) [<u>17</u>] and post-infarction cardiosclerosis. Hypertension status was ascertained based on medical records and current use of antihypertensive medication at enrollment. Diagnosis of IHD was established according to the World Health Organization criteria. All patients underwent diagnostic evaluation including exercise stress testing and/or coronary angiography, assessment of chest pain symptoms, electrocardiography (ECG) and measurement of cardiac biomarkers (troponin, myoglobin and creatine kinase). Blood pressure was measured at recruitment using standard procedures and systolic (SBP)/diastolic (DBP) blood pressure values were recorded for all participants.

Exclusion criteria were conditions that could confound inflammatory or metabolic status including diabetes, inflammatory diseases, local inflammatory processes, allergic reactions, signs of renal insufficiency, circulatory insufficiency of the stage III, Alzheimer’s disease, presence of hyperthermia and insolation during the previous 2-3 weeks.

Control blood samples were obtained from 230 healthy donors without clinical manifestations of IHD and arterial hypertension, without family history of premature atherosclerosis or ischemic events at ECG, oncology, autoimmune diseases, known hereditary diseases and acute/chronic inflammatory diseases. Controls were selected to match the patient group based on personal information collected via questionnaires.

All participants completed a structured questionnaire capturing ethnicity, demographic characteristics, lifestyle factors (tobacco use, alcohol consumption, diet) and medical history.

### Genomic DNA isolation

DNA was extracted from EDTA-treated peripheral blood samples by using «ReliaPrep™ Blood gDNA Miniprep System» (*Promega, USA).* Qualitative and quantitative characteristics of the DNA samples were estimated by spectrophotometry.

### Genotyping and quality control

Study participants were genotyped using the Illumina Global Screening Array-24 v3.0 Bead Chip (Illumina Inc., San Diego, California, U.S.). Genotype calling and initial clustering were performed using the GenomeStudio Genotyping Module v2.0. Genotype data were exported in Illumina A/B allele format together with marker annotations, including rsID, chromosome and genomic position. Sample quality control (QC) included assessment of individual genotype call rate, sex concordance and duplicate/replicate concordance. Samples with the call rate below 0.95, sex discordance or unresolved duplicate status were excluded from the downstream analysis. SNP level QC included filtering by SNP call rate, minor allele frequency and Hardy-Weinberg equilibrium in controls. SNPs with the call rate below 0.98 were excluded. SNPs with minor allele frequency below 0.03 were excluded during variant level QC. SNPs deviating from the Hardy-Weinberg equilibrium (HWE) in controls at P < 1 × 10^-6^ were excluded. GWAS analysis was restricted to autosomal variants and sex chromosome, XY region variants were excluded because they require chromosome specific genotype coding and sex-aware association modelling.

### Statistical analysis

#### Clinical statistics

Baseline clinical characteristics were summarized as mean ± standard deviation (SD) for continuous variables and as counts (percentage) for categorical variables. Comparisons between *case* and *control* groups were performed using Student’s t-test for independent samples for continuous variables including age and systolic/diastolic blood pressure. The presence of arterial hypertension was considered as a categorical variable and compared between groups using the chi-square test or Fisher’s exact test as appropriate. All statistical tests were two-sided.

#### Linkage disequilibrium pruning and SNP selection for principal component analysis

Genotype data were initially obtained in allele-based format from Illumina Genome Studio 2.0, together with SNPs annotations including chromosome, base pair position and marker summary information. Before population structure analysis, SNP preprocessing steps were performed in Python using pandas and NumPy.

First, SNPs located within known long-range linkage disequilibrium (LD) regions were excluded. These regions are characterized by the extended LD patterns that may disproportionately influence principal component analysis (PCA) and obscure genome-wide population structure. Long-range LD regions were defined according to previously published recommendations for the GRCh37/hg19 genome build (Price et al., 2008). These regions included the major histocompatibility complex (MHC) on chromosome 6 (25-35 Mb) and other established high-LD intervals across autosomes. SNPs falling withing these intervals were removed based on their chromosomal coordinates, while genotype and annotation columns were preserved. After removal of long-range LD regions, genotypes were converted from allele-based calls to numeric dosage values coded as 0, 1 or 2 representing the number of minor alleles per SNP. Minor and major alleles were inferred empirically withing the study sample for each SNP. Missing or non-informative genotype calls were encoded as missing values.

Genome-wide LD pruning was then performed on the autosomal dosage data using a custom sliding-window approach analogous to the PLINK—indep-pairwise procedure(Purcell et al., 2007) (Chang et al., 2015). Withing each chromosome, SNPs were ordered by genomic position and evaluated in windows of 50 SNPs advanced by 5 SNPs at each step. Pairwise LD between SNPs was quantified using the squared Pearson correlation coefficient (r^2^) computed from the available dosage values. For SNP pairs with r^2^>0.2 one SNP from the correlated pair was removed according to the pruning procedure. This process was iteratively applied across windows and chromosomes to obtain an approximately LD-independent set of autosomal markers for principal component analysis, consistent with standard PCA-based population structure workflows in genetic association studies (Price et al., 2006a) (Patterson et al., 2006) (Price et al., 2006b).

#### Principal component analysis

Population structure was evaluated by the principal component analysis (PCA) using the LD-reduced set of autosomal SNPs obtained after exclusion of long-range LD regions and genome-wide LD pruning. The pruned genotype dataset was represented as minor-allele dosage values and read into Python. Metadata columns were removed leaving a genotype matrix with SNPs as rows and samples as columns. The matrix was then transposed to obtain a [samples × SNPs] matrix for the PCA.

Before decomposition, missing genotype values were imputed using the SNP specific mean dosage. Genotypes were then standardized per SNP by subtracting the mean dosage and dividing by the sample standard deviation. SNPs with standard deviation equal to zero were assigned a standard deviation of one to avoid division by zero. PCA was computed using singular value decomposition (SVD) of the standardized matrix with reduced SVD (Harris et al., 2020a) (Patterson et al., 2006). Principal component scores for each individual were obtained from the left singular vectors scaled by the singular values and the first ten principal components (PC1 − PC10) were retained. The resulting per-sample PC score table was exported for downstream visualization (PC1 vs PC2 scatterplots) and for inclusion of PC1 − PC10 as covariates for downstream association analysis (Price et al., 2006b). To reduce the influence of population outliers, one outlier-removal iteration was conducted based on the initial PCA results. Samples located outside the main cluster in PC1 − PC2 sparse were excluded and PCA was recomputed on the remaining individuals. The recomputed PC1 – PC10 scores were used for final visualization and as covariates in downstream association analysis.

#### Genome-wide association analysis

Genome-wide association analysis was performed in Python 3.12.12 using pandas (version 3.0.0) (The pandas development team, 2026), NumPy (version 2.4.1) (Harris et al., 2020b) and statsmodels (version 0.14.6) (Seabold and Perktold, 2010). Association testing was conducted by logistic regression under an additive genetic model. For each SNP, the genotype dosage was coded as 0, 1 or 2, corresponding to the number of Illumina B alleles. Disease status was used as the dependent variable with cases coded as 1 and controls coded as 0. Age, sex and the first ten principal components were included as covariates to adjust for demographic effects and population structure.

For each variant the following logistic regression model was fitted:

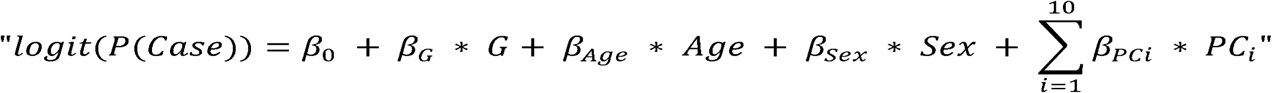

where G represents the SNP genotype dosage coded as the number of Illumina B alleles, Age is the age at the examination, Sex is a binary covariate and PC1 − PC10 are the first ten principal components.

Association testing was performed using the logit function implemented in the statsmodels Python library. The SNP effect was assessed using the regression coefficient for the B allele dosage term. Odds ratios (ORs) were calculated as the exponential of the regression coefficient (*β*G) and therefore represented the per B-allele effect. Corresponding 95% confidence intervals (95 % CI) were derived from the standard errors of the estimated genotype coefficients. Variants for which logistic regression failed to converge or produced unstable estimates were reported with missing association statistics. For each SNP, allele frequencies of the Illumina A and B alleles were calculated and minor allele frequency was defined as *MAF = min (f_A_, f_B_)*. Thus, MAF was used as a marker level frequency measure and did not necessarily correspond to the effect allele (B allele) frequency. Illumina A/B allele codes were converted to nucleotide alleles using the GSA-24v3-0_A1 Strand Report annotation file for annotation of selected top-ranked variants and for preparation of the complete GWAS summary statistics file for repository submission. The final GWAS summary statistics were converted to the GWAS-SSF-compatible fields. Variants with invalid nucleotide allele annotation or inconsistencies between GWAS summary statistics and genotype-derived sample size or allele frequency checks were excluded from the repository file. Duplicate and conflicting duplicate variant records were removed retaining one record per unique autosomal variant key.

GWAS results were visualized using Manhattan and quantile-quantile (QQ) plots based on SNP association p-values. Manhattan plots displayed −log10(p) values across genomic coordinates with horizontal lines indicating the Bonferroni-corrected significance threshold and an exploratory suggestive threshold. The Bonferroni-corrected threshold was calculated as 0.05 divided by the number of SNPs tested. QQ plots were used to assess calibration of association statistics and potential test statistic inflation. The genomic inflation factor (λ_GC_) was calculated as the median of the observed chi-square statistics divided by the expected median of a chi-square distribution with one degree of freedom.

### Knowledge graph-guided reanalysis of GWAS summary statistics

To complement the single-SNP GWAS, we reanalysed the association summary statistics using a knowledge graph-guided framework based on heterogenous graph neural networks (KGWAS) (Huang et al., 2024a). KGWAS is designed to improve discovery in small cohort GWAS by integrating GWAS summary statistics with a large functional genomics knowledge graph across variants and genes. The official KGWAS implementation provides an API for loading the knowledge graph, importing external GWAS summary statistics, processing the GWAS file, preparing train/validation/test splits and training the graph neural network model.

#### GWAS summary statistics used as an input

The primary GWAS summary statistics were obtained using logistic regression adjusted for sex, age and the first ten genetic principal components and corresponded to the per B allele effect as described above. For each SNP, the summary statistics included rsID, chromosome, base-pair position (GRCh37), genotype-effect regression coefficient (β), standard error (SE), odds ratio (OR), 95% confidence interval (95% CI), p-value, minor allele frequency and total sample size (N). For KGWAS input, a tab-delimited summary statistics file (sumstats.tsv) was prepared including rsID, chromosome, position, beta, SE, 95 % CI, p-value, MAF and sample size. Additional association statistics were retained when required by the implementation. The file did not include nucleotide alleles columns because downstream analysis focused on SNP significance and gene prioritization rather than on comparison of allelic effect directions with external GWAS datasets. Variants with missing or non-finite values, invalid p-values or missing rsIDs were removed. Only SNPs that could be matched by rsID to variant nodes in the KGWAS knowledge graph were retained for downstream analysis.

#### Knowledge graph and KGWAS model training

KGWAS analysis was performed in a Miniforge conda environment using Python 3.12.12. The official KGWAS Python implementation was used with the precompiled heterogeneous knowledge graph and default model settings, except for the specified number of training epochs and batch size (NumPy Developers, 2026). The graph comprises three main node types: 1) variant nodes initialized with sequence-based embeddings, derived from the Enformer regulatory sequence model (Avsec et al., 2021); 2) gene nodes initialized with protein sequence embeddings from ESM-type protein language models (Rives et al., 2021); 3) Gene Ontology (GO) term nodes connected to genes and variants via functional edges. After loading the knowledge graph, the external GWAS summary statistics were imported and mapped to KGWAS variant nodes by rsID. The intersecting set of SNPs present both in the GWAS summary statistics and in the KGWAS graph was used for model training and prediction.

A heterogeneous graph neural network was trained to model the GWAS signal using the graph structure and node embeddings. We used the default KGWAS model architecture while training was performed for 5 epochs with a mini-batch size of 256. Model performance was monitored on the validation set using mean squared error and the Pearson correlation between observed and predicted GWAS signals. After training, KGWAS produced model derived SNP-level scores and exported them in a MAGMA compatible format (KGWAS_magma_format_∗.csv), including SNP identifiers, genomic coordinates and KGWAS-derived association statistics.

### Conventional and KGWAS-informed MAGMA gene level analysis

Gene level association analysis was performed using MAGMA v1.10 (de Leeuw et al., 2015a). Two complementary MAGMA based analyses were conducted. First, conventional MAGMA was applied to the original GWAS SNP p-values. Second, KGWAS-informed MAGMA was applied to the KGWAS derived SNP p-values generated from the same GWAS summary statistics. This design allowed comparison between gene level aggregation based directly on the observed GWAS evidence and gene level prioritization based on the KGWAS reweighted SNP signals. For both analyses, MAGMA was run in SNP-wise mean mode using SNP p-value input file. To generate the SNP p-value input file, the final autosomal GWAS summary statistics were cross-checked for SNP identifier consistency and filtered to retain one record per rsID. Duplicate rsID records were removed before export resulting in the final input file sumstats_final_for_MAGMA_KGWAS.tsv, which contained SNP identifiers, p-values and sample size. In this mode, MAGMA uses the supplied SNP p-values together with an LD reference panel to aggregate SNP evidence into gene level statistics. The genotype data supplied through –bfile was used as LD reference data. MAGMA applies the SNP-wise mean model for gene analysis when SNP p-values are provided through –pval.

SNPs were mapped to genes using the gene annotation file distributed with KGWAS based on genomic position. The annotation step assigns SNPs to genes according to their genomic coordinates. All analyses were conducted using GRCh37/hg19 coordinates to ensure consistency between GWAS summary statistics, gene annotation and LD reference data. MAGMA reference data from the 1000 Genomes Project phase 3 are provided in Build 37 coordinates which is consistent with this genome build.

To evaluate sensitivity to LD reference structure, MAGMA analyses were performed using multiple LD reference panels. The in-sample Kazakh reference panel was used as the primary population matched LD reference. This reference was generated from the post-QC genotype dataset converted to PLINK binary format and was used only for LD estimation in MAGMA. In addition, 1000 Genomes phase 3 East Asian (EAS), European (EUR) and South Asian (SAS) reference panels were used as ancestry sensitivity analyses (Auton et al., 2015). The SAS panel was retained as an additional exploratory non-European Asian reference panel. For each LD reference setting, MAGMA was run with the corresponding PLINK binary reference files, SNP p-value file through --pval and gene annotation file specified through --gene-annot. For conventional MAGMA, the input consisted of the original GWAS SNP p-values. For KGWAS-informed MAGMA the input files consisted of KGWAS SNP association statistics in MAGMA compatible format. The same gene annotation and LD reference settings were used for conventional MAGMA and KGWAS-informed MAGMA to ensure comparability between the two approaches. MAGMA generated output files containing chromosome, gene start and end positions, number of mapped SNPs, effective number of parameters, sample size and genes p-values. The results files were imported into Python for post-processing procedures. For each gene we computed the midpoint of the annotated interval and used it as the genomic coordinate for plotting, with −log_10_(P-value) of the gene on the y-axis. Genes were ordered by chromosome and position to construct genome wide Manhattan plots at the gene level. A Bonferroni corrected significance threshold was defined as 0.05 divided by the number of tested genes and the corresponding −log10(a/M) line was overlaid on the plots. Lower “suggestive” threshold (p < 1×10^−4^) was additionally depicted on the plot. Entrez Gene IDs from MAGMA were mapped to the HUGO Gene Nomenclature Committee (HGNC) symbols and full gene names using an external annotation service (MyGene.info) and these annotations were used in the gene tables and for labeling top genes in the figures (Wu et al., 2013).

## Results

### Cohort characteristics

Initially, 509 individuals were recruited and genotyped. After sample quality control, 39 individuals were excluded because of sex discordance, duplicate checks, PCA outlier status or missing essential covariate information. Detailed sample quality-control summary results are provided in Supplementary Table S1. The final GWAS dataset included 470 individuals comprising of 240 cases with IHD and co-occurring arterial hypertension and 230 apparently healthy controls. Baseline characteristics of the final GWAS cohort are shown in Table 1.

**Table 1.**
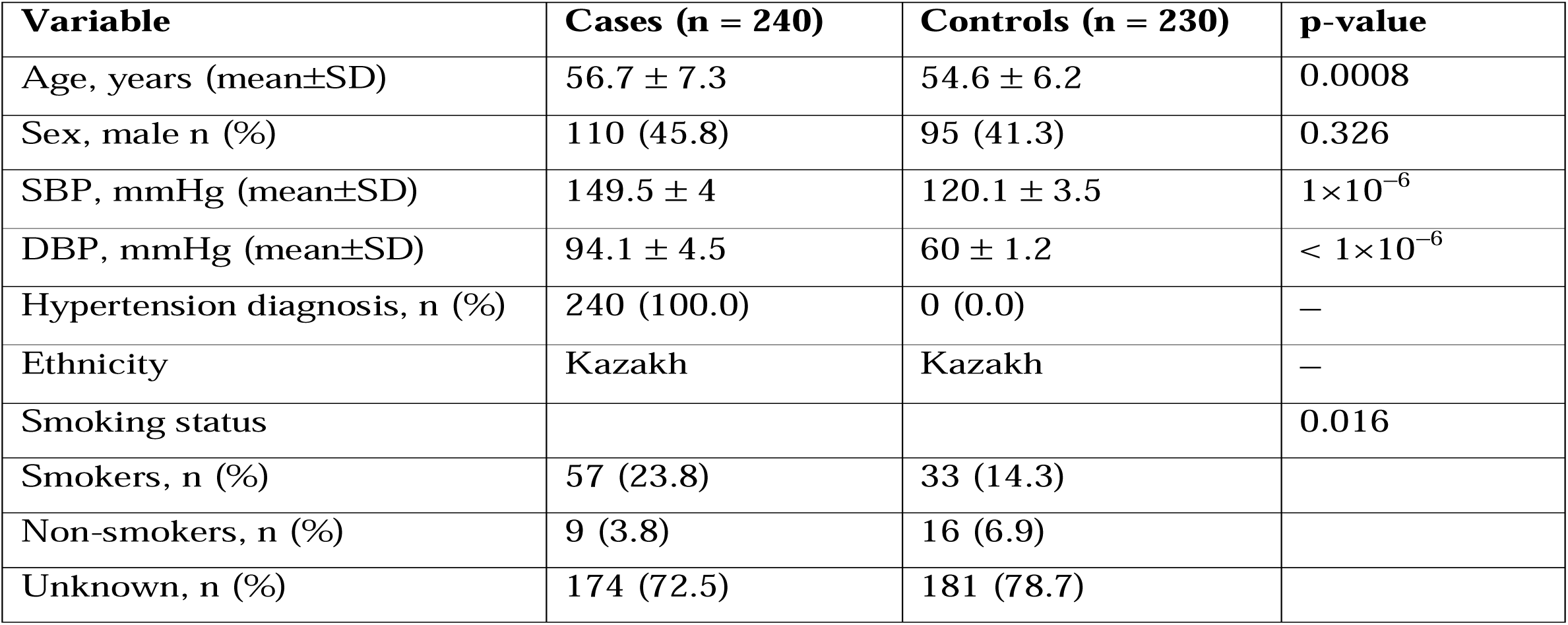
Baseline characteristics of the final GWAS cohort after sample quality control.

| Variable | Cases (n = 240) | Controls (n = 230) | p-value |
| --- | --- | --- | --- |
| Age, years (mean±SD) | 56.7 ± 7.3 | 54.6 ± 6.2 | 0.0008 |
| Sex, male n (%) | 110 (45.8) | 95 (41.3) | 0.326 |
| SBP, mmHg (mean±SD) | 149.5 ± 4 | 120.1 ± 3.5 | $1 \times 10^{-6}$ |
| DBP, mmHg (mean±SD) | 94.1 ± 4.5 | 60 ± 1.2 | $< 1 \times 10^{-6}$ |
| Hypertension diagnosis, n (%) | 240 (100.0) | 0 (0.0) | – |
| Ethnicity | Kazakh | Kazakh | – |
| Smoking status |  |  | 0.016 |
| Smokers, n (%) | 57 (23.8) | 33 (14.3) |  |
| Non-smokers, n (%) | 9 (3.8) | 16 (6.9) |  |
| Unknown, n (%) | 174 (72.5) | 181 (78.7) |  |

Cases were statistically older than controls, although the absolute difference was modest (56.7 ± 7.3 years in cases and 54.6 ± 6.2 years in controls, p = 0.0008, Cohen’s d = 0.31). No significant difference in sex distribution was observed between cases and controls with males comprising 45.8% and 41.3% of the groups, respectively (p = 0.326)

As expected, blood pressure values were markedly higher in the cases than in the controls consistent with the case definition that included comorbid arterial hypertension. Mean systolic blood pressure (SBP) was 149.5 ± 4 mmHg in cases and 120.1 ± 3.5 mmHg in controls (p < 1×10^-6^) and mean diastolic blood pressure (DBP) was 94.1 ± 4.5 mmHg in cases and 60 ± 1.2 mmHg in controls (p < 1×10^-6^). All participants self-identified as Kazakh. Smoking information was incomplete in both groups. Smoking status (smokers, non-smokers, unknown) distribution differed between cases and controls (χ^2^ test, p = 0.016). Among participants with known smoking status, a higher proportion of smokers was observed in the cases (57/240, 23.8%) than in the controls (33/230, 14.3%). However, the majority of participants did not report their smoking status (unknown: 174/240, 72.5% in the cases and 181/230, 78.7% in the controls) limiting the conclusions based on self-reported smoking categories.

### Genome-wide association analysis

After genotype re-clustering in a single GenomeStudio project using the GSA v3 annotation files, followed by sample- and variant- level QC, the final GWAS dataset comprised 470 individuals including 240 cases and 230 controls. Detailed variants quality-control summary results are provided in Supplementary Table S1. Association testing was performed using logistic regression under an additive genetic model adjusted for age, sex and the first ten principal components. Smoking was not included as a covariate in the primary model because of substantial missingness in self-reported smoking status.

A total of 325 976 post QC autosomal variants entered the association analysis of which 325 976 variants had valid regression statistics and were retained for downstream interpretation. Using the Bonferroni correction for multiple testing the genome-wide significance threshold was set to P < 1.53 × 10^-7^. No variant reached this threshold in the final analysis (Figure 1). The strongest SNP association was observed for rs1511924 at chromosome 16:73,805,209 mapped to *ZFHX3* gene (effect allele C; OR = 0.49, 95% CI 0.37–0.66; p = 1.00 × 10^-6^), showing a protective direction of the effect. Additional top-ranked variants included rs6009715 at chromosome 22:49,351,462 (effect allele G; OR = 1.80, 95% CI 1.37–2.36; P = 2.30 × 10^-5^), rs66598464 at chromosome 4:100,323,833 near *ADH1C/ADH17* gene (effect allele G; OR = 2.13, 95% CI 1.49–3.02; P = 2.75 × 10^-5^), rs711516 at chromosome 7:109,374,419 (effect allele C; OR = 2.19, 95% CI 1.52–3.17; P = 2.95 × 10^-5^) and rs11814051 at chromosome 10:121,283,241 within *RGS10* gene (effect allele C; OR = 0.37, 95% CI 0.23–0.60; P = 2.98 × 10^-5^) (Table 2). The QQ plot demonstrated close agreement between observed and expected p-value distributions with a genomic inflation factor of λGC = 1.046, indicating adequate control for residual population stratification and model inflation.

**Figure 1.**
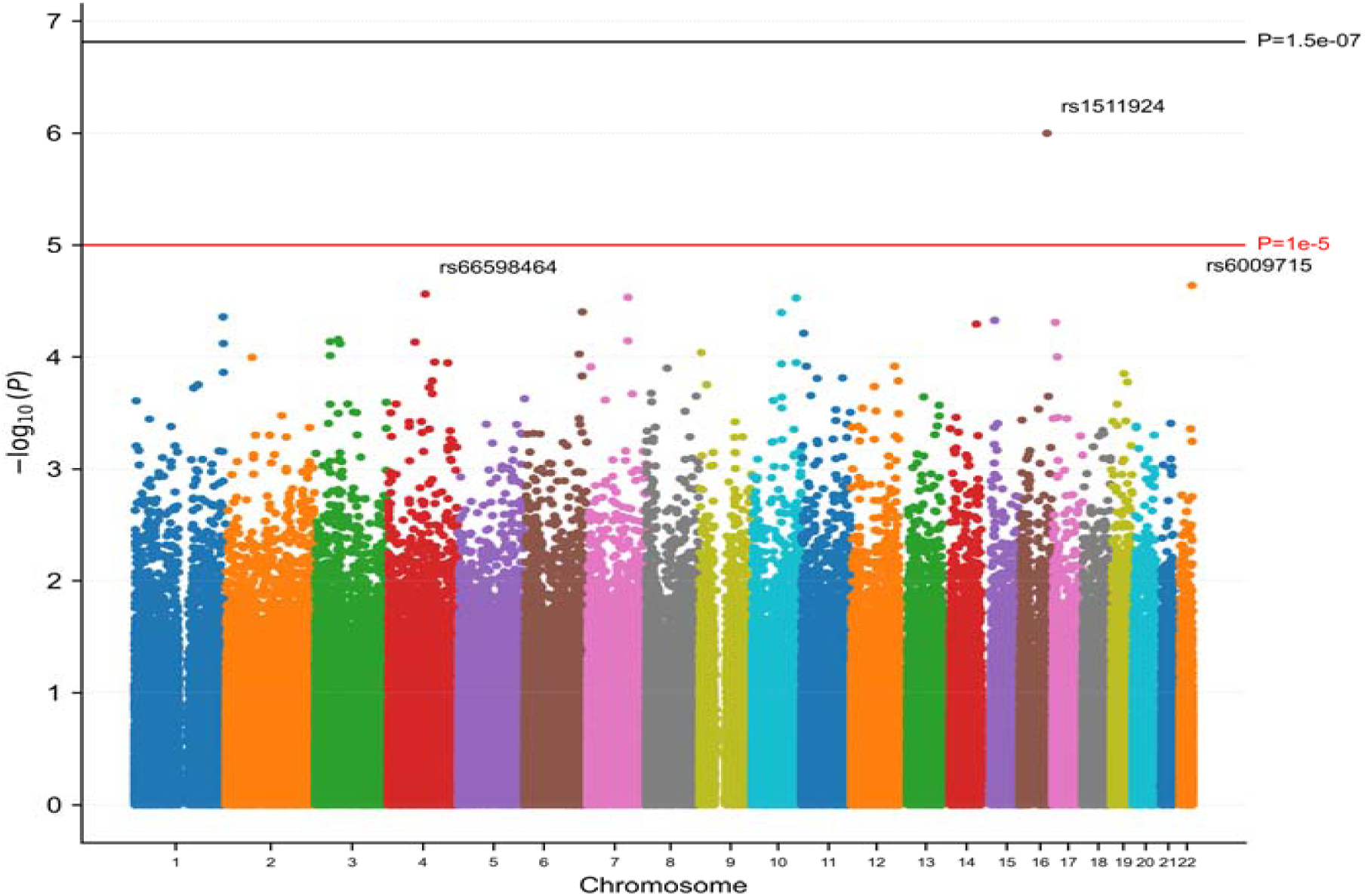
Manhattan plot of the GWAS results for IHD with co-occurring arterial hypertension. Genome-wide association results from logistic regression under an additive genetic model adjusted for age, sex and first ten principal components (PC1 − PC10). Each point represents an SNP plotted by chromosomal position with the y-axis showing −log_10_(P). The horizontal black line indicates the Bonferroni-corrected genome-wide significance threshold (p = 1.53×10^-7^) and the red line indicates a suggestive threshold (p = 1×10^-5^). The most significant variants are annotated by the rsID.

**Table 2.** Top associated SNP variants from the GWAS.

| RsID | Chr | Position | Nearest gene | Distance to gene, bp | Effect allele | Other allele | OR for the effect allele | 95% CI | p-value | EAF | MAF |
| --- | --- | --- | --- | --- | --- | --- | --- | --- | --- | --- | --- |
| rs1511924 | 16 | 73805209 | ZFHX3 | 0 | C | A | 0.495 | 0.37 - 0.656 | 1.00E-06 | 0.573 | 0.426 |
| rs6009715 | 22 | 49351462 | - | - | G | A | 1.802 | 1.37 - 2.36 | 2.30E-05 | 0.559 | 0.441 |
| rs66598464 | 4 | 100323833 | ADH1C;<br>ADH17;<br>ENSG00000299279<br>lncRNA | 32666 | G | A | 2.126 | 1.49 - 3.02 | 2.75E-05 | 0.230 | 0.230 |
| rs711516 | 7 | 109374419 | ENSG00000308019<br>lncRNA | 0 | C | T | 2.193 | 1.52 - 3.17 | 2.95E-05 | 0.194 | 0.194 |
| rs11814051 | 10 | 121283241 | RGS10 | 0 | C | T | 0.375 | 0.23 - 0.6 | 2.98E-05 | 0.885 | 0.115 |
| rs9383776 | 6 | 156142928 | ENSG00000287092<br>lncRNA | 0 | G | T | 1.748 | 1.33 - 2.29 | 3.96E-05 | 0.519 | 0.481 |
| rs164235 | 10 | 80329628 | - | - | C | T | 0.345 | 0.21 - 0.57 | 4.03E-05 | 0.096 | 0.096 |
| rs12038803 | 1 | 241522325 | RGS7;<br>ENSG00000286496.<br>2 lncRNA | 1795 bp<br>upstream; 0 | G | A | 0.501 | 0.36 - 0.7 | 4.40E-05 | 0.778 | 0.222 |
| rs76313777 | 15 | 34490450 | KATNBL1 | 0 | C | T | 2.171 | 1.5 - 3.14 | 4.70E-05 | 0.173 | 0.173 |
| rs4465647 | 17 | 7244477 | ACAP1 | 0 | G | A | 2.161 | 1.49 - 3.13 | 4.90E-05 | 0.170 | 0.171 |

Abbreviations: EAF – effect allele frequency; MAF – minor allele frequency

For public summary-statistics deposition, additional allele annotation checks, genotype derived sample size and allele frequency consistency checks and duplicate variant removal were applied resulting in 325 495 unique autosomal variant records in the GWAS Catalog submission file.

### Gene-based analyses using conventional MAGMA and KGWAS-informed MAGMA

As secondary exploratory gene-level analyses, we performed conventional MAGMA analysis directly using SNP-level p-values from our GWAS summary statistics file and, separately, KGWAS-informed MAGMA analysis using KGWAS-derived SNP-level scores. The final autosomal GWAS summary statistics file included 325 976 variants and was processed into the MAGMA compatible input file containing 325 686 unique SNPs after duplicate rsID removal. Given the admixed Eurasian genetic structure of the Kazakh population and the absence of a large public Kazakh-specific LD reference panel, we used an in-sample Kazakh LD reference as the primary reference for the gene-based analyses. EAS and EUR 1000 Genomes panels were used as continental ancestry sensitivity references, while SAS was included as an additional exploratory non-European Asian reference panel.

Conventional MAGMA analysis did not identify genes reaching the Bonferroni-corrected significance threshold across the evaluated LD reference panels (Figure 2). The strongest signals were observed for *BMAL1* and *SMCO4* genes, both of which were consistently ranked among the top genes across EAS, EUR, SAS and KAZ LD references. *BMAL1* gene showed p-value of 3.38 × 10^-4^, 3.92 × 10^-4^ and 3.12 × 10^-4^ in the EAS, EUR and KAZ analyses, respectively. Similarly, *SMCO4* gene showed p-value of 3.68 × 10^-4^, 5.42 × 10^-4^, 5.42 × 10^-4^ and 3.35 × 10^-4^ across the same reference panels. Other prioritized genes included *GSTM3*, *PRSS58*, *BICD2*, *SHDL2*, *KATNBL* and *OR6T1*, although several of these signals were sensitive to the LD reference panel used (see Supplementary Table 2). For example, *GSTM3* gene ranked first in the SAS analysis and third in the KAZ analysis but was substantially lower ranked in the EUR analysis.

**Figure 2.**
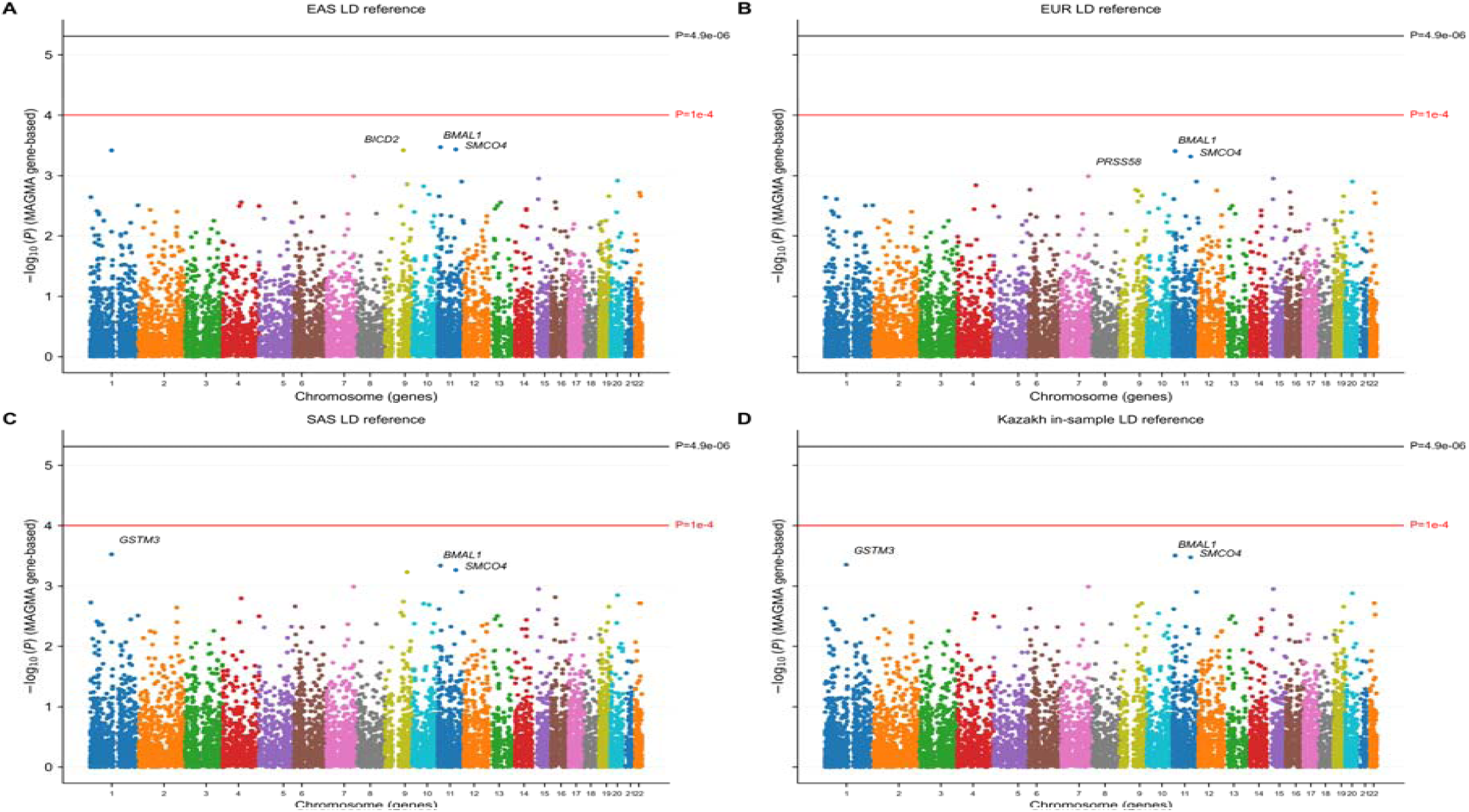
Gene-based Manhattan plots from the MAGMA analysis. **(A)** Results using the 1000 Genomes East Asian (EAS) LD reference panel; (B) Results using the 1000 Genomes European (EUR) LD reference panel; (C) Results using the 1000 Genomes South Asian (SAS) LD reference panel; (D) Results using the Kazakh in-sample LD reference panel. Each point represents a gene positioned at the midpoint of its annotated genomic interval and colored by chromosome. The y-axis shows −log_10_ of the MAGMA gene-based p-value. The upper horizontal line indicates the Bonferroni-corrected significance threshold (P ≈ 4.9 × 10^-6^). The horizontal red line marks the suggestive threshold (P = 1 × 10^-4^).

In contrast, KGWAS-informed MAGMA produced a more pronounced and more focused set of gene-level signals (Figure 3, Table 3). The Bonferroni-corrected threshold for KGWAS-informed MAGMA was approximately 4.8–4.9 × 10^-6^ depending on the number of genes tested in each LD-reference analysis. Under this threshold, *TLE1* gene reached significance across all four LD reference panels making it the most stable KGWAS-informed gene level signal. *TLE1* gene ranked first across EAS, EUR, KAZ and SAS analyses with p-values 1.77 × 10^-6^, 1.72 × 10^-7^, 1.76 × 10^-6^ and 1.03 × 10^-6^, respectively. *CSMD1* gene was also consistently prioritized ranking second in the EAS, EUR, SAS analyses and third in the KAZ analysis. However, *CSMD1* gene reached the Bonferroni significance threshold in the EAS, EUR and SAS but not in the in-sample KAZ reference panel. *SMCO4* gene ranked among the top three genes across all four KGWAS-informed MAGMA analyses and exceeded the Bonferroni threshold in the EAS and KAZ panels with p-values of 3.92 × 10^-6^ and 3.36 × 10^-6^, respectively. In the EUR and SAS analyses, *SMCO4* gene showed slightly weaker but still highly ranked signals with p-values of 6.30 × 10^-6^, 7.55 × 10^-6^, respectively. This indicates a reference sensitive but recurrent KGWAS-informed signal. Additional sub-threshold or suggestive signals included *HSDL2*, *C6orf10*, *GSTM3* and *PCDH15* genes.

**Figure 3.**
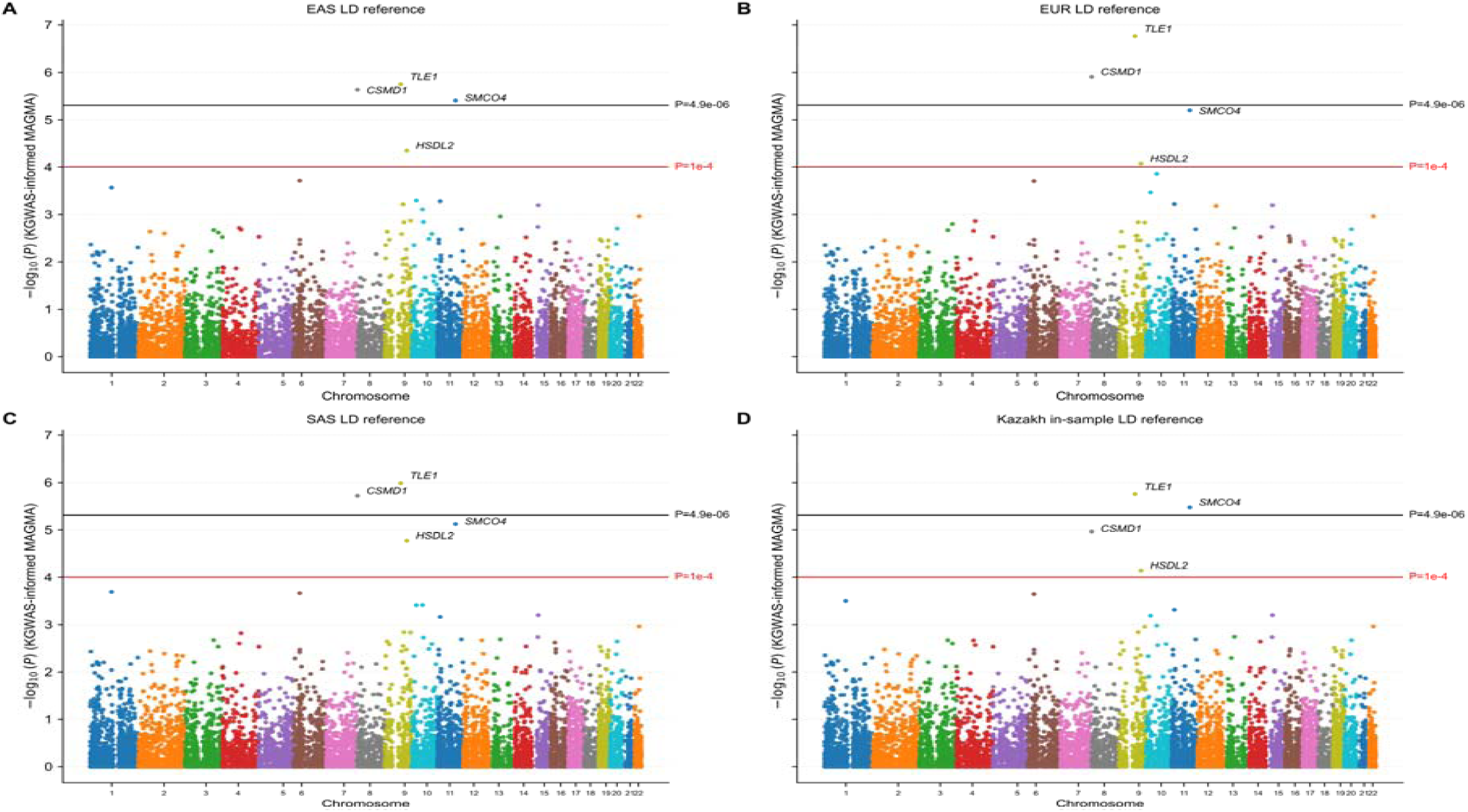
Gene-based Manhattan plots from the KGWAS-informed MAGMA analysis. **(A)** Results using the 1000 Genomes East Asian (EAS) LD reference panel; (B) Results using the 1000 Genomes European (EUR) LD reference panel; (C) Results using the 1000 Genomes South Asian (SAS) LD reference panel; (D) Results using the Kazakh in-sample LD reference panel. Each point represents a gene positioned at the midpoint of its annotated genomic interval and colored by chromosome. The y-axis shows −log_10_ of the MAGMA gene-based p-value. The upper horizontal line indicates the Bonferroni-corrected genome-wide significance threshold (P ≈ 4.9 × 10^-6^). The horizontal red line marks the suggestive threshold (P = 1 × 10^-4^).

**Table 3.**
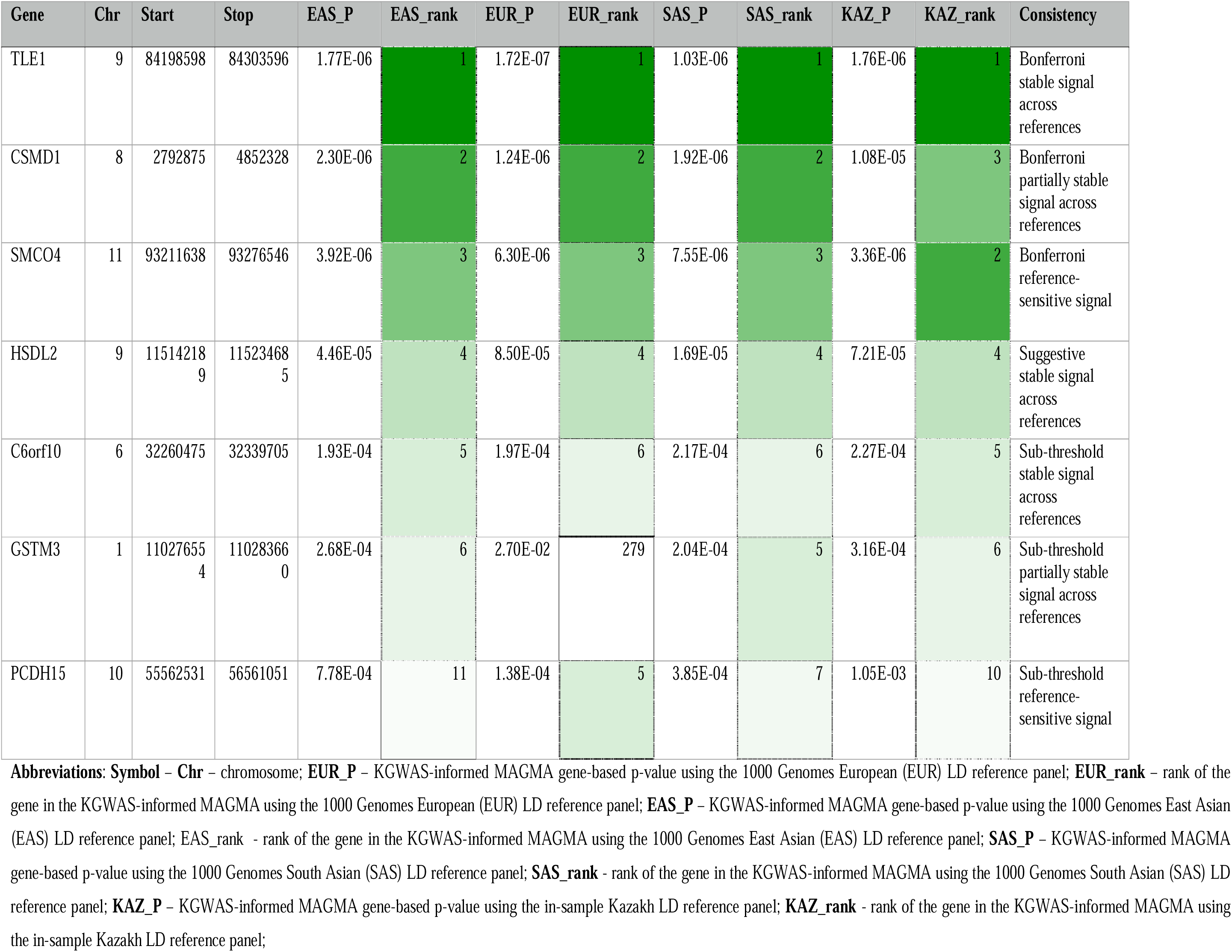
Stable KGWAS-informed MAGMA gene-level candidates across four LD-reference panels.

Comparison of the conventional MAGMA and KGWAS-informed MAGMA results showed partial overlap between the two analytical strategies. *SMCO4* gene was consistently prioritized in both analyses, although the KGWAS-informed MAGMA analysis strengthened its gene level signal. Notably, results of the standalone KGWAS without subsequent MAGMA analysis, showed rs11601368 mapped to *SMCO4* gene as a top-ranked SNP signal. The standalone KGWAS SNP results are summarized in Supplementary Table 3. *GSTM3* and *HSDL2* genes also appeared among the prioritized genes in both conventional MAGMA and KGWAS-informed MAGMA frameworks but their evidence remained sensitive to the reference panel or sub-threshold. By contrast, *TLE1* and *CSMD1* genes became substantially stronger in the KGWAS-informed MAGMA analysis than in the conventional MAGMA suggesting that KGWAS reweighting increased the contribution of SNP level evidence within these regions.

To better understand the origin of the KGWAS-informed gene level signals, we examined variant level results within selected prioritized gene regions. Supplementary Table 4 summarizes 165 SNPs mapped to the *CSMD1*, *TLE1* and *SMCO4* genes including their original GWAS p-values and KGWAS derived p-values. In the *CSMD1* gene, 157 variants were evaluated; 12 had nominal GWAS evidence (P<0.05), whereas 19 had KGWAS derived *P* values below 0.05, including 9 variants that were not nominally significant in the original GWAS. In *TLE1* gene, 3 of 5 variants had nominal GWAS support, whereas all 5 variants showed KGWAS derived *P* < 0.05. In the *SMCO4* gene, 2 of 3 variants were nominally associated in the original GWAS and all 3 had KGWAS derived P < 0.05. These findings indicate that the KGWAS-informed MAGMA results were driven by model-based reweighting of SNP level evidence within the selected genes regions rather than by the conventional GWAS significance alone.

Importantly, although the *CSMD1* gene showed one of the strongest KGWAS-informed MAGMA signals at the gene level, it is a large gene represented by many variants most of which did not show nominal association in the original GWAS. In contrast, *TLE1* gene showed a more compact and consistent pattern of support with all mapped variants strengthened by KGWAS and reaching KGWAS derived P < 0.05. This makes the *TLE1* signal the most consistent candidate in terms of variant level prioritization whereas the *CSMD1* signal may be more influenced by the gene size and the aggregation of multiple weak variant level effects across a broad genomic interval.

In summary, this pilot GWAS of IHD-HTN co-occurrence phenotype in the Kazakh cohort did not identify SNP associations that passed the Bonferroni-corrected significance threshold. However, the strongest SNP signal mapped to the *ZFHX3* locus, a region with prior cardiovascular relevance. Conventional MAGMA analysis did not identify Bonferroni significant gene level associations consistent with the limited power of the cohort studied. In contrast, KGWAS-informed MAGMA showed stronger candidate gene prioritization. The most stable signal was obtained for the *TLE1* gene across the LD reference panels studied, including in-sample Kazakh LD panel. The *SMCO4* gene was supported by both conventional MAGMA and KGWAS-informed MAGMA in the Kazakh LD reference analysis with the KGWAS-informed reweighting and strengthening the signal to the Bonferroni significant level. The *CSMD1* gene reached the Bonferroni significance for the ancestry related EAS and EUR reference panels but this signal was not supported when the in-sample Kazakh LD reference was used. Together, these results demonstrate the feasibility of genome-wide and knowledge graph-guided analyses in this under-studied population and provide hypothesis-generating targets for the replication and functional validation.

## Discussion

In the present study, we performed a case-control GWAS of IHD-HTN co-occurrence phenotype in the Kazakh cohort using logistic regression adjusted for age, sex and population structure. The final SNP analysis did not identify any variant surpassing the Bonferroni-corrected significance threshold. Nevertheless, the QQ plot showed close agreement between observed and expected P-value distributions and the genomic inflation factor was low (λGC = 1.046), indicating adequate calibration of the association statistics and no evidence of substantial residual inflation.

The absence of genome-wide significant SNP associations is not unexpected given the modest sample size and the complex nature of the IHD-HTN co-occurrence phenotype. IHD with co-occurrence of arterial hypertension is likely influenced by multiple biological pathways, including vascular remodeling, blood pressure regulation, lipid and metabolic processes, inflammation, endothelial function and environmental exposures. Therefore, the present GWAS should be interpreted as a pilot discovery analysis rather than a definitive locus discovery study.

The strongest SNP signal was observed for the rs1511924 at chromosome 16 mapped to the *ZFHX3* gene. This locus has prior cardiovascular relevance: variant rs7193343-T has been associated with atrial fibrillation and ischemic/cardioembolic stroke in a combined analysis of stroke samples (Gudbjartsson et al., 2009). Another variant in this locus rs2106261 has been reported as an atrial fibrillation susceptibility in European ancestry cohorts and in Chinese Han populations (Li et al., 2011). Additional *ZFHX3* variants including rs6499600 and rs16971436 have also been investigated in relation to atrial fibrillation in Chinese Han populations with rs6499600 and rs2106261 showing significant associations and rs16971436 showing borderline evidence (Liu et al., 2014). Although our rs1511924 signal did not reach the Bonferroni-significance threshold, the prior findings at the *ZFHX3* locus supports its consideration as hypothesis generating cardiovascular candidate region for Kazakh population that requires an independent replication. Other SNP signals, also, did not yield Bonferroni significant loci and motivate to use of gene level approaches that may capture aggregated or biologically informed signals not evident at the levels of individual variants. We therefore compared the conventional MAGMA gene testing with the KGWAS-informed MAGMA, in which the KGWAS-derived SNP statistics were used for downstream gene prioritization.

The comparison between conventional MAGMA and KGWAS-informed MAGMA highlights both the potential and the limitations of gene level prioritization in the small Kazakh cohort. Conventional MAGMA analysis directly aggregates SNP GWAS evidence within genes while accounting for local LD structure and therefore remains closely tied to the observed association statistics from the present cohort. In our analysis, conventional MAGMA did not identify any genes reaching the Bonferroni-corrected significance threshold across the evaluated LD reference panels, which is consistent with the absence of genome-wide significant SNP-level associations and the limited statistical power of the cohorts. Nevertheless, several genes, including the *BMAL1*, *SMCO4*, *GSTM3* and *HSDL2*, showed nominal or reference-dependent evidence indicating the presence of weak gene level aggregation signals that did not reach corrected significance. MAGMA was originally developed to improve gene and gene-set analysis by aggregating SNP level signals while accounting for LD and multi-marker effects but it remains dependent on the strength and quality of the underlying GWAS summary statistics (de Leeuw et al., 2015b).

In contrast, KGWAS-informed MAGMA produced stronger and more focused gene level prioritization, particularly for the *TLE1*, *CSMD1* and *SMCO4* genes. This pattern is consistent with the intended purpose of KGWAS which is designed to improve discovery in small-cohort GWAS by integrating GWAS summary statistics with a large functional genomics knowledge graph across variants and genes (Huang et al., 2024b). However, this also changes the interpretation of the resulting gene level statistics. Unlike conventional MAGMA, KGWAS-informed MAGMA does not aggregate only the original GWAS p-values. It uses KGWAS-derived SNP level scores that incorporate model-based information and external functional genomic priors. Therefore, KGWAS-informed MAGMA results should be interpreted as model-informed gene prioritization rather than as independent statistical confirmation of gene-level association.

Among the KGWAS-informed MAGMA prioritized genes, *TLE1* gene appeared to be the most stable candidate. It ranked first across all four LD reference panels and reached the Bonferroni-corrected threshold in each analysis. A more detailed view of the underlying SNPs in this gene showed a coherent pattern: among the five variants mapped to the *TLE1* gene, three had nominal support in the original GWAS, whereas all five had KGWAS-derived P values below 0.05. This suggests that KGWAS strengthened a compact set of weak but locally consistent SNP signals. In contrast, *CSMD1* gene requires more cautions interpretation. Although it was strongly prioritized in the KGWAS-informed MAGMA and reached Bonferroni-corrected significance in the EAS, EUR and SAS analyses, it did not reach the corrected threshold in our in-sample KAZ reference analysis. Moreover, *CSMD1* gene is a large gene represented by many mapped variants. In our detailed SNP analysis, only 12 of 157 variants showed nominal GWAS evidence, while 19 had prioritized by KGWAS with p-values below 0.05. Thus, the *CSMD1* signal may be influenced by the gene size and the aggregation of multiple weak SNP signals across its broad genomic interval. This distinction is important because the physical location of a variant within a gene does not necessarily imply that the variant functionally affects the same gene. This is particularly relevant for large genes as *CSMD1* where many mapped variants are likely to be intronic or non-coding. Non-coding variants may regulate nearby or distal genes, tag causal variants through linkage disequilibrium or be prioritized through broader graph connectivity rather than direct functional action on gene in which they are located. Therefore, the KGWAS-informed MAGMA signal for the *CSMD1* gene should be interpreted as a gene-region or locus-level prioritization rather than as direct evidence that the mapped variants functionally affect *CSMD1* gene itself. Functional assignment of non-coding variants would require additional evidence such as tissue-relevant eQTL, chromatin interaction, enhancer annotation, fine-mapping or colocalization analyses. The interpretation of the KGWAS-informed results also requires distinction between variant level and gene level evidence. In the present workflow, KGWAS first reweighted SNP-level association statistics using functional genomic information whereas gene-level prioritization was generated in the subsequent MAGMA step by mapping and aggregating KGWAS SNP-level scores within genes regions while accounting for LD. This distinction is important because KGWAS SNP-level signals may become stronger after model-based reweighting but the final gene-level ranking depends on how these reweighted variants are assigned to genes and aggregated by MAGMA. This was particularly evident for *SMCO4* gene. In addition to being one of the strongest nominal signals in conventional MAGMA analysis, *SMCO4* gene also contained the strongest variant-level KGWAS signal among the prioritized loci examined. The SNP rs11601368 in *SMCO4* gene had nominal support in the original GWAS (P = 3.69 × 10^-4^; β = - 0.81; OR = 0.45; MAF = 0.115; N = 470) and its signal was further strengthened after KGWAS reweighting with a KGWAS P value of 2.29 × 10^-5^. After downstream MAGMA aggregation of KGWAS SNP level evidence, *SMCO4* gene reached the Bonferroni-significant threshold in the EAS and our in-sample KAZ reference panels. Thus, *SMCO4* gene represents a recurrent but reference sensitive signal supported at both the conventional MAGMA and KGWAS variants levels.

Our findings also align with emerging methodological concerns regarding knowledge-graph-based GWAS approaches. Recent work on context-aware KGWAS has emphasized that the original KGWAS framework relies on a large general-purpose knowledge graph. This may introduce spurious correlations and may lack sufficient resolution for a specific disease context. This work further suggests that cell-type specific or context dependent knowledge graphs may improve biological robustness and interpretations of the disease pathways development (Jiang et al., 2026). In this context, the present results illustrate a key interpretational challenge: KGWAS may strengthen weak SNP signals in underpowered GWAS by incorporating external functional genomic priors but the subsequent gene-level prioritization depends on MAGMA based SNP to gene mapping, LD structure of populations analyzed and aggregation of KGWAS scores.

Therefore, when the primary GWAS evidence is limited KGWAS-informed MAGMA results may partly reflect model-driven reweighting and broad functional connectivity rather than disease specific genetic architecture alone.

The use of multiple LD reference panels further supports the need for the ancestry sensitivity analyses. *TLE1* gene remained significant across all reference settings suggesting a relatively stable KGWAS-informed MAGMA signal. However, prioritization results of the *CSMD1* and *SMCO4* genes showed dependence on reference panels analyzed. *CSMD1* gene failed to reach the Bonferroni thresholds in our in-sample KAZ reference and *SMCO4* gene reached the threshold only in the EAS and KAZ panels. This variability is important in the context of the admixed Eurasian genetic structure of the Kazakh population. The in-sample KAZ reference panel is likely the most relevant approximation of LD structure for the present cohort, whereas EAS and EUR serve as continental ancestry sensitivity reference. The SAS 1000 Genomes panel was retained only as an additional exploratory non-European Asian reference panel.

Overall, these results support a cautious interpretation of KGWAS-informed MAGMA gene prioritization in small cohorts. *TLE1* gene may represent the most consistent KGWAS-informed MAGMA candidate because it showed a stable gene level significance across LD references and coherent SNP level support in the GWAS. *SMCO4* gene is also notable because it was supported by the conventional MAGMA analysis, standalone KGWAS SNP level evidence and KGWAS-informed MAGMA analysis for the in-sample Kaz reference. *CSMD1* gene remains an important candidate gene-locus signal but its interpretation should account for the gene size, the predominance of non-coding mapped variants and the possibility that KGWAS reweighting amplified dispersed weak signals across the broad locus. Thus, KGWAS-informed MAGMA results in this study should be considered as hypothesis-generating and useful for prioritizing candidates for the future replication and functional validation rather than definitive evidence of causal gene-level involvement.

## Limitations

This study has several limitations that should be considered when interpreting the results. First, the sample size of the Kazakh cohort was modest for the SNP level GWAS, especially for a complex co-occurrence phenotype such as IHD-HTN. As a result, the study had limited statistical power to detect variants with small or moderate effects and the absence of Bonferroni significant SNP associations should not be interpreted as evidence of no genetic contribution to the phenotype. Second, the analysis was performed in a single Kazakh cohort without an independent replication dataset. Therefore, the identified SNPs, genes and locus signals, such as *ZFHX3*, *TLE1*, *SMCO4* and *CSMD1*, should be considered hypothesis-generating until confirmed in larger and independent Kazakh or Central Asian cohorts.

Third, as mentioned above, the IHD-HTN phenotype is clinically and biologically heterogeneous. Ischemic heart disease and hypertension may share some genetic and environmental determinants but they may also arise through partially distinct mechanisms. The present case-control design may not fully capture differences in disease duration, severity, medication use, lifestyle factors, metabolic status or other clinical covariates that could influence genetic associations. Fourth, although population structure was adjusted using principal components and the genomic inflation factor indicated adequate calibration, residual bias associated with the mixed Eurasian ancestry of the Kazakh population cannot be completely excluded.

Fifth, interpretation of the gene level analyses depends on the statistical framework and LD reference panel used. Conventional MAGMA analysis remains dependent on the strength of the original GWAS summary statistics, whereas KGWAS-informed MAGMA incorporates model derived SNP reweighting based on external functional genomic information. Therefore, KGWAS-informed MAGMA results should not be interpreted as independent confirmation of association but rather as model-informed prioritization. In addition, differences across KAZ, EAS, EUR LD reference panels indicate that some signals are ancestry and reference sensitive. The in-sample KAZ LD panel is likely the most appropriate reference for the present cohort. The other panels should be interpreted as sensitivity analyses rather than direct substitutes for the specific Kazakh LD structure.

Sixth, MAGMA gene mapping according to the GWAS SNP scores is based primarily on genomic position and does not necessarily establish functional relationships between variants and the genes to which they are assigned. This limitation is particularly relevant for large genes such as *CSMD1* gene, where many mapped variants may be intronic or non-coding and may regulate other nearby or distal genes through regulatory mechanisms. Finally, this study did not include functional validation, fine-mapping, eQTL, chromatin interaction or tissue-specific regulatory analyses. Such analyses are needed to determine whether prioritized variants and genes have direct biological relevance to vascular, cardiac, endothelial or blood pressure related pathways.

## Conclusions

In conclusion, this pilot GWAS of the IHD-HTN co-occurrence phenotype in the Kazakh cohort did not identify SNP associations reaching the Bonferroni-corrected significance threshold. Nevertheless, the GWAS statistics were well calibrated, indicating that the analysis provided a reliable basis for the downstream exploratory prioritization despite limited statistical power.

The main contribution of the present study is not definitive locus discovery but the evaluation of the SNP GWAS, conventional MAGMA and KGWAS-informed MAGMA as complementary approaches for analyzing a complex cardiovascular co-occurrence phenotype in the underpowered and understudied population specific cohort. Conventional MAGMA analysis remained closely dependent on the original GWAS signals and did not reveal significant gene level associations, whereas KGWAS-informed MAGMA produced more focused candidate prioritization by incorporating external functional genomic information. This comparison demonstrates both the potential and the interpretational limitations of knowledge-graph-informed prioritization in a small cohort GWAS. Among the prioritized signals, *TLE1* gene showed the most stable KGWAS-informed MAGMA evidence across LD reference panels, while *SMCO4* gene represented a recurrent but reference sensitive candidate gene supported by both conventional and KGWAS-informed analyses for our in-sample Kazakh reference panel. *CSMD1* gene should be interpreted more cautiously as a broad locus signal rather than direct evidence of causal gene involvement. Thus, the study demonstrates that gene level and knowledge-graph-informed approaches may help extract biologically interpretable candidate signals from underpowered GWAS but they cannot substitute for adequate statistical power, replication and functional validation.

## AI-assisted language editing statement

The authors used an AI-assisted tool to support English language translation and editing. All content was reviewed and approved by the authors who take full responsibility for the accuracy and integrity of the manuscript.

## Data availability

Genome-wide association summary statistics generated in this study have been submitted to the NHGRI-EBI GWAS Catalog under the accession GCST90841470. Supplementary tables are provided with this article. Individual genotype and phenotype data are not publicly available because the informed consent and ethics approvals do not permit open sharing of individual genetic data. Additional analysis files and scripts are available from the corresponding author upon reasonable requests, subject to institutional and ethical restrictions.

## Data Availability

All data produced are available online at the NHGRI-EBI GWAS Catalog under the accession GCST90841470

## Acknowledgments

We thank all patients and volunteers for the participation in this study.

## Author contributions

Conceptualization: Liliya Skvortsova. Formal analysis: Liliya Skvortsova.

Investigation: Liliya Skvortsova, Kanagat Yergali, Asel Zhaksylykova, Mamura Begmanova.

Methodology: Liliya Skvortsova, Almagul Mansharipova. Supervision: Liliya Skvortsova.

Visualisation: Liliya Skvortsova, Kanagat Yergali. Writing – original draft: Liliya Skvortsova.

Writing – review and editing: Almagul Mansharipova.

